# Multimodal deep learning enhances genomic risk prediction for cardiometabolic diseases in UK Biobank

**DOI:** 10.1101/2025.04.28.25326564

**Authors:** Taiyu Zhu, Upamanyu Ghose, Héctor Climente-González, Joanna M. M. Howson, Sile Hu, Alejo Nevado-Holgado

## Abstract

Cardiometabolic diseases are multifactorial disorders influenced by numerous genetic variants and their complex interactions. Although recent studies have advanced the understanding of genetic risk prediction, current approaches predominantly rely on linear models that may not fully capture the complex, non-linear relationships between genetic factors. Here, we present DeepGP (Deep learning-based Genome-wide Predictor), a novel multimodal deep learning framework that incorporates bidirectional state space modules to predict cardiometabolic disease risk using genome-wide variants and demographic data. We conducted extensive experiments to evaluate DeepGP’s performance. First, in simulation studies incorporating joint genetic and environmental interactions, we demonstrated DeepGP’s superior prediction performance across varying levels of heritability. When evaluated on eight cardiometabolic diseases in European ancestry cohorts from the UK Biobank, DeepGP achieved significantly higher accuracy compared with conventional polygenic risk scores and machine learning methods. Model interpretability analysis identified both well-established genes and potential new signals contributing to the risk of the disease. Further validation on populations with African and Caribbean ancestries showed robust transferability of the model. Our results demonstrate the potential of cutting-edge deep learning technologies to enhance risk stratification for complex diseases across diverse ancestries and to improve disease understanding.

## Main

Cardiometabolic diseases (CMDs), comprising a wide spectrum of cardiovascular and metabolic disorders, impose a substantial global burden due to their high rates of morbidity and mortality. The alarming growth of CMD prevalence highlights the critical need for practical and effective strategies focused on early detection and prevention [1]. Many CMDs are polygenic, with many genetic variants across the genome conferring disease risk. Therefore, human genetic approaches have emerged as powerful tools in the discovery of new targets for CMDs by identifying genome-wide causal variants and disease risk prediction by aggregating the causal genetic effects of genome-wide variants into a predictive score. In recent decades, genome-wide association studies (GWAS) [2] have emerged as one of the most important human genetic approaches. GWAS have enabled the identification of numerous genetic variants, typically single nucleotide polymorphisms (SNPs), associated with various phenotypes, thus offering insights into their heritability and underlying biology. Applying GWAS in extensive study cohorts is essential to improve the estimation of genetic effects by maximizing available samples. The UK Biobank is a valuable resource that exemplifies this scale, with more than 500,000 participants in the United Kingdom [3], offering deep genetic and phenotypic data and allowing researchers to conduct large-scale studies, such as GWAS of rare variants for hundreds of traits and diseases [4, 5].

One of the most important applications emerging from GWAS is polygenic risk scores (PRS) [6]. Conventionally, these scores predict disease risk by aggregating a large number of SNPs across the genome, even though many of them may have minor effects, as a weighted sum score, with the weight of each variant estimated from external GWAS studies. This approach leverages the cumulative impact of numerous genetic variants, enabling a more comprehensive assessment of an individual’s genetic risk for a given disease. As the number of variants included in the PRS increases, the predictive power typically enhances significantly [6]. Therefore, incorporating a broader array of genetic variants, such as those from whole genome, into genetic prediction offers significant potential for early intervention and personalized medicine approaches, whilst the interpretation for the disease risk from PRS requires careful consideration and caution. Several recent studies have utilized PRS to predict CMDs in different large-scale biobank datasets [7–15], including the UK Biobank. However, traditional GWAS approaches rely on linear or logistic regression models under the assumption of additive effect to correlate genetic variants with phenotypes. Recent analysis of the UK Biobank data has revealed significant non-additive genetic effects that remain largely unexplored in conventional GWAS frameworks [16]. Since effect size estimations in these models are primarily marginal, they do not fully address the heterogeneous patterns of linkage disequilibrium (LD) across ancestry groups or capture complex non-linear effects, such as gene-by-gene or gene-by-environment interactions. As a result, PRS-based methods frequently face issues with lack of portability, especially when LD structures in populations and genetic architectures are different, limiting their predictive accuracy and utility across diverse ancestries [17]. Alternatively, machine learning algorithms have demonstrated remarkable power in further identifying and prioritizing SNPs with small effects, surpassing the capabilities of traditional statistical methods [18–20]. For instance, light gradient-boosting machines (LGBM) [21] have emerged as a powerful approach, proving effective for handling high-dimensional genomic and proteomic data [22–24]. Deep learning [25] has also demonstrated great potential for tackling complex problems in genomics [26–29], which stacks multiple layers of artificial neurons to learn intricate patterns and representations from raw data, achieving state-of-the-art performance in various tasks. Deep neural networks, particularly convolutional neural networks (CNNs), have demonstrated remarkable success in predicting the effects of genetic variants [30–32]. Transformer-based architectures [33] which are the backbones of most large language models, have been successfully adapted to predict gene expression by treating DNA sequences similarly to sentences in human languages [34, 35]. Several pioneering studies have attempted to use deep learning to map the complex relationships between genotypes and phenotypes for complex diseases, such as Alzheimer’s disease [36–38], amyotrophic lateral sclerosis [29, 39], and Parkinson’s disease [29]. However, applying these methods to whole-genome studies presents significant challenges. The high dimensionality of genetic data, which contains millions of variants, creates substantial computational constraints, making genome-wide analysis difficult with traditional neural network architectures. Furthermore, there remains a critical research gap in exploring these models for CMDs and in validating their performance across diverse ancestries.

To address these limitations, we propose DeepGP (Deep learning-based Genome-wide Predictor), a multimodal deep learning framework that enhances CMD risk prediction by modeling both additive and non-additive genetic effects across the genome. Unlike previous approaches that struggle with computational demands, DeepGP implements Mamba [40], a selective state space model (SSM) that achieves comparable performance to Transformers while maintaining linear computational scaling. The SSM architecture has recently established itself as a new paradigm for foundation models across various domains, including language processing [40], computer vision [41], and DNA sequencing [42]. DeepGP leverages this architecture by incorporating bidirectional SSM (Bi-SSM) modules to process genome-wide SNPs from each chromosome separately while integrating additional information from environmental factors from the UK Biobank. This approach outperforms traditional machine learning, deep learning, and PRS methods in our experiments. The superior performance enables the development of robust and generalizable genomic risk prediction models for CMDs.

## Results

### Overview of Methods and Workflow

Fig. 1 illustrates the overview of the data processing pipeline and the proposed deep learning framework. As shown in Fig. 1a, our methodological pipeline consists of several key steps. In order to control for effects of genetic ancestry, participants were first categorized into European (*n* = 364, 541), African, and Caribbean cohorts based on their self-reported ancestry and their projection onto the principal components (PCs). Then the process begins with SNP quality control (QC) using procedures from an existing pipeline [43]. This comprehensive QC process includes batch-level control and genotype filtering based on specific criteria: a minimum allele frequency threshold of 0.01, maximum missing genotype rate of 0.1, and Hardy-Weinberg equilibrium test *p*-value threshold of 1 × 10^*−*5^. During this stage, we also exclude related participants to minimize potential confounding effects from cryptic relatedness. Based on the kinship coefficient, we remove up to 3^*rd*^ degree relative. The SNP data is extracted from the imputed genotype data in the UK Biobank [3].

**Fig. 1:**
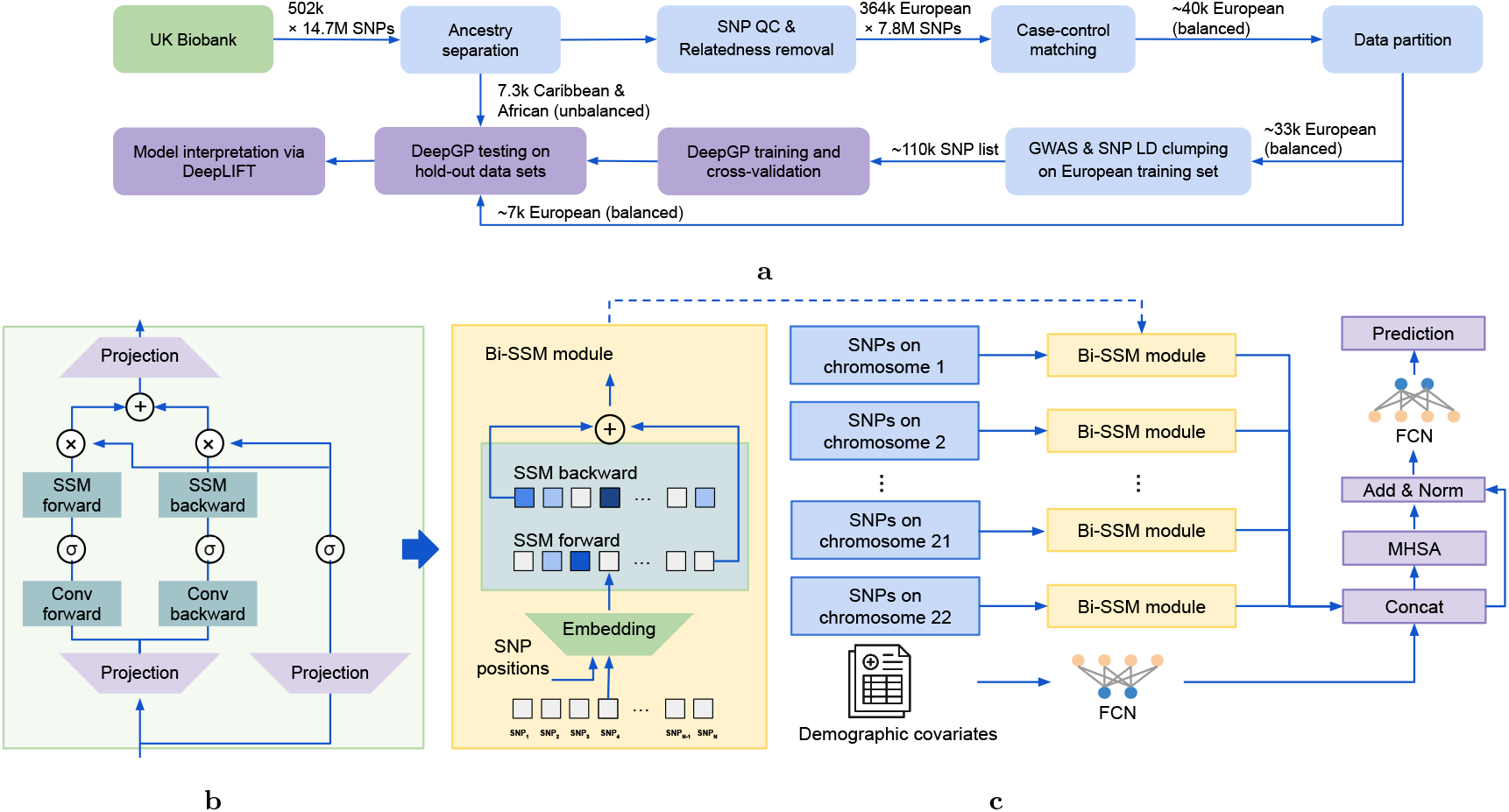
Overview of the proposed disease risk prediction model. **a**, Data preprocessing pipeline to extract genotype profiles from the UK Biobank. The input data includes imputed genotype data, and covariates reflecting environmental effect, including sex, age, and six principal components (PCs) of population structure. **b**, Architecture of a Bi-SSM, which consists of bidirectional selection mechanism to capture the complex interactions and long-range dependencies between SNPs within each chromosome. *σ* is a non-linear activation function. **c**, Architecture of DeepGP for predicting the risk of CMDs. The SNP data from each of the 22 autosomal chromosomes is processed by separate Bi-SSM modules, while the covariates are processed by a fully connected network (FCN) branch. The outputs from these parallel processing paths are then integrated through a lightweight multi-head self-attention (MHSA) module and a final FCN layer to generate risk predictions.

Thereafter, we perform case-control matching to create balanced European training and testing sets, while retaining the diverse ancestral groups to assess the model’s generalizability and transferability. Subsequently, we perform GWAS and corresponding LD clumping in European training sets to select the independent leading SNP within each LD block. This step reduces the effect of genetic linkage, which removes variants with correlated effects to maximize information, and decreases the dimensionality of input data to accommodate memory constraints of computational hardware.

We combine this processed SNP data with demographic covariates, including sex, age, and six PCs of population ancestry from the UK Biobank to jointly fit the model by accounting for effects of genetic, environmental and their interactions. In addition, the inclusion of PCs helps control for potential confounding effects of population stratification.

DeepGP is then trained, cross-validated, and evaluated on both the hold-out testing set and two validation cohorts. Finally, we perform model interpretability analysis using DeepLIFT [44], which computes feature importance scores through backpropagation of activation. This analysis identifies the most informative SNPs and explores non-linear interactions, providing insights into the model’s decision-making process.

To effectively learn hidden patterns from SNP data, a Bi-SSM is employed to extract hidden features in both forward and backward directions (Fig. 1b), inspired by the success of bidirectional recurrent neural networks in sequence data analysis [45]. Bi-SSM modules are applied to SNP data on each of the 22 autosomal chromosomes (Fig. 1c), allowing the model to capture chromosome-specific patterns and interactions. The covariates are processed by a separate fully-connected network (FCN). Then the hidden representations are aggregated through concatenation and further processed by output layers, including a multi-head self-attention (MHSA) module, to make the final predictions.

### Predictive Performance in Simulation Studies

We generated synthetic phenotypes by modeling joint genetic, environment, gene-by-gene interaction effects, and gene-by-environment interactions effects based on the imputed genotype data, which enables modeling comprehensive scenarios of varying genetic architectures and interaction patterns. We then evaluated the performance of DeepGP and other commonly used baseline methods, including PRS and LGBM, on these synthetic datasets and made comparisons.

Specifically, we used LD-clumped SNPs from the type 2 diabetes data set and maintained the same balanced European training and test set partitioning (Supplementary Table 3), and followed the training, cross-validation and evaluation procedures as shown in Fig. 1a. In particular, the phenotype was simulated as a combination of genetic effects, gene-gene interactions, gene-environment interactions and a random error due to unobserved environment factors. After proper rescaling, the variance of the phenotype was set as 1. The variance of the genetic effects is equivalent to the SNP heritability 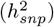. We further set variance of both gene-gene and gene-environment interactions as fixed 0.2, and varied 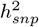 from 0.1 to 0.6 to assess model performance across different heritability levels. For each 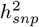 value, we performed 10 independent simulations using different random seeds to ensure robust evaluation of model performance.

Fig. 2 illustrates the area under the receiver operating characteristic curve (AUROC) for each method across different simulated scenarios. To ensure a robust evaluation, we repeated the simulation of phenotypes 10 times for each scenario, using different random seeds. All three methods (DeepGP, LGBM, and PRS) were trained and tested on identical datasets, with input of both SNPs and covariates, during each simulation. Statistical significance of performance differences was evaluated using *t*-tests or Mann-Whitney U tests [46], depending on the normality of the difference distributions of AUROC.

**Fig. 2:**
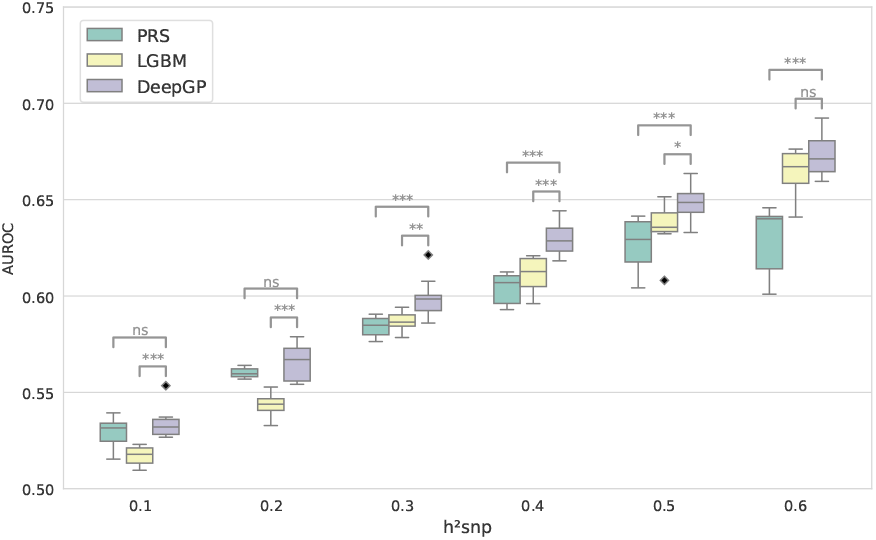
Predictive performance comparison across SNP heritability thresholds. AUROC for DeepGP, LGBM, and standard PRS plotted against varying levels of SNP heritability 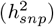. Diamond markers represent outliers. Statistical significance: ^***^*p* < 0.001, ^**^*p* < 0.01, ^*^*p* < 0.05.

DeepGP consistently outperforms the baseline methods across all SNP heritability settings. The most notable improvements are observed when SNP heritability ranges from 0.3 to 0.5, where DeepGP demonstrates a statistically significant advantage over both LGBM and PRS. As heritability increases to 0.6, the performance gap between DeepGP and LGBM narrows. These results demonstrate DeepGP’s superior predictive ability by capturing complex genetic architectures, particularly in scenarios with moderate heritability where traditional methods may struggle to fully model the intricate relationships between genetic variants and phenotypes.

### Enhanced Risk Prediction for Cardiometabolic Diseases

We evaluated DeepGP and baseline methods for predicting eight CMDs in the UK Biobank: atherosclerosis, atrial fibrillation, cardiovascular disease, coronary artery disease, heart failure, myocardial infarction, stroke, and type 2 diabetes. For fair evaluation, we used balanced hold-out testing data with equal numbers of cases and controls (Supplementary Table 3).

Table 1 presents the accuracy and AUROC results for DeepGP, LGBM, and PRS. DeepGP consistently achieved the highest accuracy and AUROC scores across all diseases compared to both baseline methods. Additional metrics including precision, sensitivity, and specificity are provided in Supplementary Table 5, where DeepGP demonstrated the best performance across all metrics for atherosclerosis and stroke. For the remaining diseases, DeepGP maintained the most balanced performance across these additional metrics. We further compared performance with a broader range of traditional machine learning and deep learning baseline methods, including CNN, dense neural networks (Dense), multilayer perceptron (MLP), random forest (RF), and SNP-level logistic regression (LR) models, as shown in Supplementary Fig. 5. Among all considered baseline approaches, LGBM and PRS demonstrated the strongest performance, which is why we selected them as the primary comparison methods.

**Table 1:** Accuracy and AUROC of genomic Risk prediction on hold-out testing data of European ancestry.

| Phenotype | Accuracy |  |  | AUROC |  |  |
| --- | --- | --- | --- | --- | --- | --- |
|  | DeepGP | LGBM | PRS | DeepGP | LGBM | PRS |
| Atherosclerosis | **68.6** | 64.7 | 65.7 | **0.75** | 0.70 | 0.70 |
| Atrial Fibrillation | **67.6** | 65.7 | 65.9 | **0.74** | 0.71 | 0.70 |
| Cardiovascular Disease | **67.1** | 63.8 | 64.4 | **0.74** | 0.69 | 0.68 |
| Coronary Artery Disease | **67.4** | 63.9 | 63.5 | **0.74** | 0.68 | 0.69 |
| Heart Failure | **67.1** | 62.8 | 62.7 | **0.72** | 0.68 | 0.66 |
| Myocardial Infarction | **69.0** | 65.8 | 62.9 | **0.76** | 0.71 | 0.67 |
| Stroke | **63.2** | 59.6 | 57.9 | **0.68** | 0.62 | 0.60 |
| Type 2 Diabetes | **60.4** | 59.5 | 57.7 | **0.64** | 0.61 | 0.60 |

The second and third most substantial improvements were observed for stroke and heart failure, with a reduction in misclassified cases of 12.6% and 11.8%, respectively, when compared with PRS. Interestingly, these three diseases have the smallest number of cases among the eight diseases studied (Supplementary Table 3), indicating that deep learning can achieve superior performance when the size of the data is limited and traditional statistical methods often struggle with statistical power to capture non-additive effects. Deep learning’s ability to model complex, non-linear relationships between genetic variants and disease risk enables it to outperform PRS in these challenging cases, highlighting its potential for improving risk prediction in diseases with limited available data.

### Model Interpretability and Genetic Insights

To investigate the contribution of each SNP towards the prediction outputs, we performed DeepLIFT, a method for interpreting deep learning models by backpropagating the contributions of all neurons in the network to every feature of the input. This approach allowed us to derive feature importance scores for each input SNP. We then compared the DeepLIFT feature importance scores with statistics of standard GWAS.

The scatter plots in Fig. 3 depict the relationships between the DeepLIFT feature importance scores and the Chi-square statistics for each SNP across the eight CMDs. The scatter plots visualize the concordance between the feature importance scores derived from the deep learning model and the statistical significance of the SNPs determined by our GWAS analysis. The vertical blue line represents the Chi-square statistic corresponding to the widely used GWAS *p*-value threshold of 5 × 10^*−*8^, which is commonly employed to identify significant associations.

**Fig. 3:**
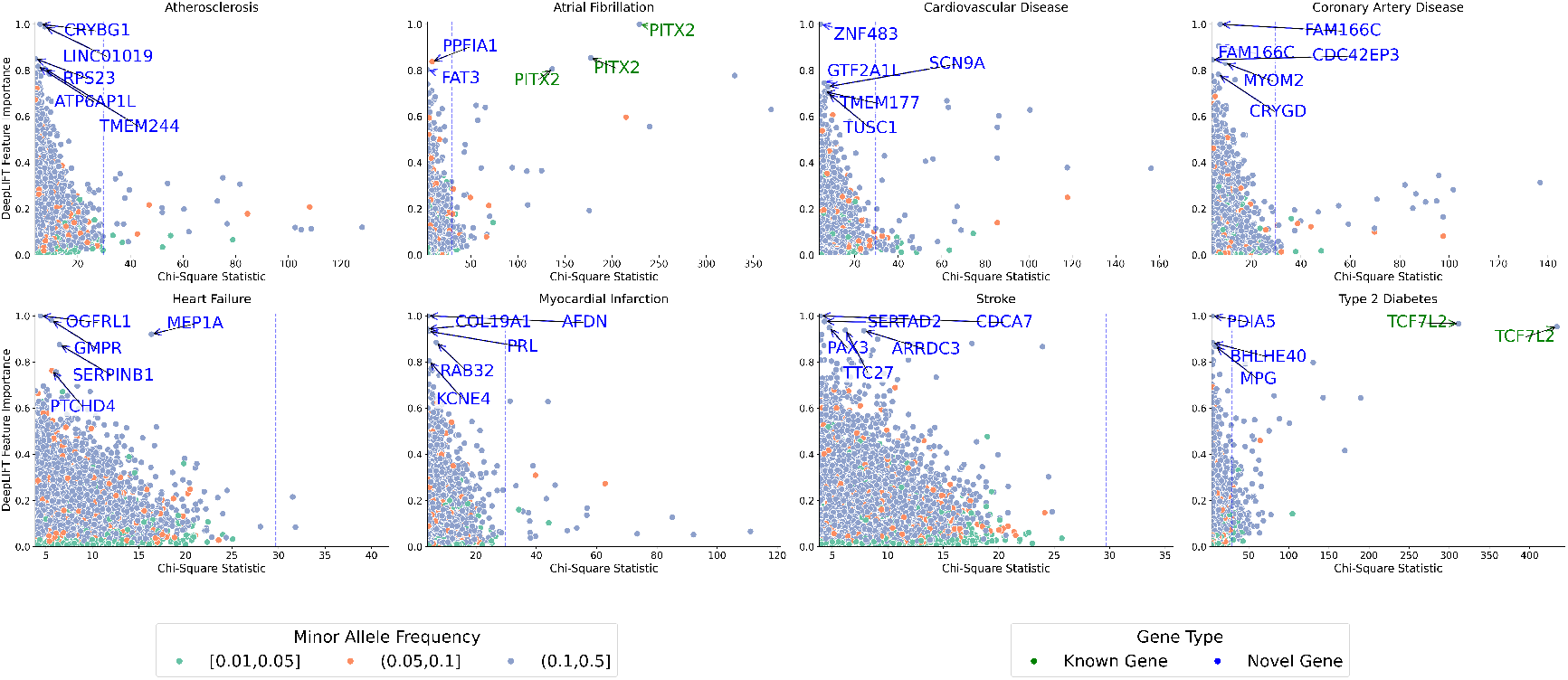
Comparison of feature importance derived from DeepLIFT and *χ*^2^ statistics for the considered CMDs. The color of each point represents the minor allele frequency, and the vertical blue line indicates the *χ*^2^ statistic corresponding to GWAS p-value threshold of 5 × 10^*−*8^. The top five SNPs with the highest feature importance scores are annotated with their nearest genes and labeled on the plots.

The genes with the most significant associations in GWAS also exhibited high feature importance scores in DeepGP. For instance, *TCF7L2* in type 2 diabetes [47] and *PITX2* in atrial fibrillation [48] have been extensively validated as key genetic risk factors across diverse populations. This observation highlights the ability of DeepGP to capture the biological relevance of these well-established genetic associations. More importantly, DeepGP identified genes in which the SNPs were not significant in conventional GWAS we conducted. For example, SNPs near *PDIA5* and *MPG*, were identified having high contribution to T2D. The direct role of *PDIA5* in diabetes related biological pathway remains unclear, but the *PDIA5* has high levels of gene and protein expression in liver and pancreas [GTEx/HPA].

Interestingly, SNPs with low minor allele frequencies between 0.01 and 0.05 did not achieve high feature importance scores, particularly for atherosclerosis, myocardial infarction, and heart failure. This finding suggests that low frequency variants may have limited contributions to the predictive performance of DeepGP for these specific diseases. This is likely due to the increased difficulty in detecting interaction effects associated with low-frequency variants, especially given the sparsity of cases for various CMDs in the UK Biobank cohort. Further investigation into the role of low frequency or even rare variants and their potential interactions with common variants could provide additional insights into the genetic architecture of these complex cardiometabolic traits [49].

### Transferability of Risk Prediction in Diverse Ancestries

DeepGP was trained on an European population from the UK Biobank. To assess its transferability, we further evaluated the model’s performance on two cohorts with diverse ethnic backgrounds: one African (*n* = 3, 143) and the other Caribbean (*n* = 4, 243), also from the UK Biobank. Fig. 4 presents the principal component analysis (PCA) plot depicting the population ancestry of these three cohorts. The PCA was used to visualize the genetic similarities and differences among the cohorts. It is worth noting that the clusters of the European training and testing sets highly overlap, indicating their genetic similarity. However, the European clusters are distinctly separated from the Caribbean population and particularly the African population, highlighting the substantial genetic dissimilarities among these groups. This observation underscores the challenge of maintaining transferability across populations with diverse ancestral backgrounds.

**Fig. 4:**
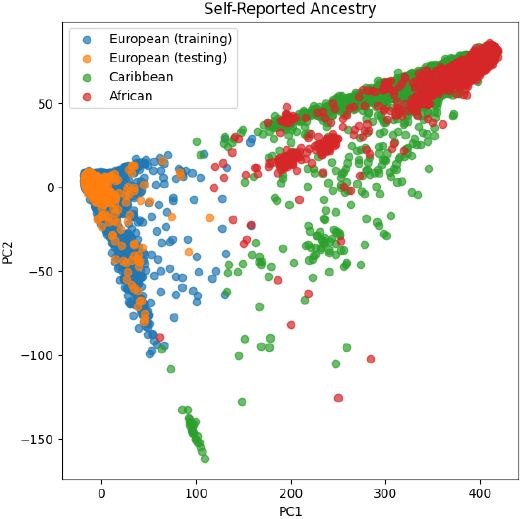
Principal component analysis to visualize the three ancestries in the UK Biobank. Two major principal components (PC1 and PC2) are used, where the color of each point represents the group of data set.

Due to the limited sample sizes of the African and Caribbean cohorts, only type 2 diabetes had more than 500 cases in both ancestries. Therefore, we evaluated the models’ performance specifically on type 2 diabetes. Given the class imbalance in the test sets, we used balanced metrics to ensure a fair assessment. These included balanced accuracy [50], AUROC, area under the precision-recall curve (AUPRC), and Matthews correlation coefficient (MCC). The results of these evaluations are summarized in Table 2. Additional evaluation metrics, including precision, sensitivity, and specificity, are provided in Supplementary Table 6. Notably, PRS exhibited poor performance in this cross-population scenario, failing to provide meaningful risk stratification. In both African and Caribbean ancestries, the PRS model’s AUROC was close to random chance (0.5), indicating a performance breakdown. This decline stems from the substantial genetic architecture differences between the European training population and the African and Caribbean test population

**Table 2:** Balanced evaluation metrics for type 2 diabetes risk prediction across African and Caribbean ancestries.

| Ancestry | Balanced Accuracy |  | AUROC |  | AUPRC |  | MCC |  |
| --- | --- | --- | --- | --- | --- | --- | --- | --- |
|  | DeepGP | LGBM | DeepGP | LGBM | DeepGP | LGBM | DeepGP | LGBM |
| African | **59.1** | 53.6 | **0.64** | 0.54 | **0.28** | 0.20 | **0.18** | 0.05 |
| Caribbean | **60.9** | 53.1 | **0.68** | 0.56 | **0.31** | 0.22 | **0.21** | 0.05 |

Meanwhile, DeepGP significantly improved the AUROC by more than 0.1 compared with the LGBM model. These results demonstrate the superior performance and transferability of DeepGP in predicting type 2 diabetes risk across populations with different ancestral backgrounds. Better performance (7/8 metrics) was observed in the Caribbean cohort compared with the African cohort, which can be attributed to the slightly closer genetic distance between European and Caribbean ancestries as visualized in Fig. 4.

## Discussion

In this study, we introduce DeepGP, a novel deep learning framework for predicting CMD risk based on genomic and demographic data. Comparative analyses against established methods, including LGBM and PRS, demonstrate the superior performance of DeepGP across both simulated phenotypes and eight real CMDs in the UK Biobank. Notably, DeepGP exhibits robust transferability when validated on populations with diverse ethnic backgrounds, addressing a critical challenge in genomic risk prediction. The consistent outperformance of DeepGP in various scenarios indicates its potential to capture complex genetic architectures and interactions, complementing the traditional PRS-based approaches.

The core module used in DeepGP is the Bi-SSM module, inspired by recent advancements in natural language processing for handling long sequence data with long-range interactions, such as DNA [42]. This choice was based on a careful consideration of model complexity, memory constraints, and efficacy. Our initial experiments explored conventional deep neural networks, including Dense and CNNs, but these models did not exhibit significant improvements compared to LGBM on prediction (Supplementary Fig. 5). We then investigated Transformer architectures, but encountered memory bottleneck on our NVIDIA A10G GPUs (24 GB) due to the quadratic time complexity and memory usage of MHSA when processing the extremely large number of SNPs. Even after performing LD clumping and dividing the input SNPs across 22 chromosomes, the maximum input length for each chromosome-specific input branch remained substantial, reaching approximately 10,000 SNP.

Consequently, Mamba emerged as the optimal choice, requiring only linear complexity while generating hidden states through selective state space scanning (see Methods for details). We further enhanced the model’s capability by implementing a bidirectional architecture that allows the model to process genetic sequences in both forward and reverse directions, enabling it to capture context and dependencies from both upstream and downstream genetic elements. Therefore, DeepGP demonstrates a superior ability of capturing long-range genetic interactions without the prohibitive memory requirements of traditional Transformer models, making it particularly well-suited for genomic risk prediction tasks.

The complexity of genetic architecture and gene-environment interaction emerge as the main challenge in successful application of statistical or machine learning methods in genetic target discovery and genetic disease prediction. Our methods leverage state-of-art deep learning method to capture additional non-additive and/or non-linear effect, including long-range gene-gene and gene-environment interaction, to identify candidate genes and refine the polygenic prediction. Although the model achieves higher prediction accuracy compared with other commonly-used methods through simulation studies, we are still able to see the potential of further improvement to our method. For instance, due to the limit of available computational resource, we did not use all the genome-wide variants to train the deep learning model, resulting in loss of information from those genome-wide rare variants and complex joint interaction involving multiple variants and environmental factors. With the increase of available whole genome sequencing data in biobank-level, improving the current model to substantially reduce the current computational burden paves a promising way to an enhancement of model prediction and portability, and a joint model by fitting genome-wide genetic variants will help uncover the endogenous aetiological insight of CMDs.

One of the most challenging problems posed by applying deep learning models in genetics is their lack of interpretability: Meaningful biological insights often hide inside the black box. To address this issue, we applied DeepLIFT to quantify the contribution from individual genes fitted in the model for the disease prediction, and prioritized them based upon their contribution. DeepLIFT successfully identified complementary candidate genes to the conventional methods. Causation of some of those identified genes were further validated by colocalisation, whilst the mechanism of causality remain unclear and is in need of further exploration. However, popular methods used for causal inference such as colocalisation and Mendelian Randomization are largely based on linear models, limiting the causal validation for non-linear effect genes identified by DeepLIFT. Finally, an improved SNP to gene (S2G) mapping strategy is essential to correctly map the variants to genes. Current S2G approaches primarily rely on existing variant annotations or, in some cases, the physical distance between the variant and the gene region. However, this method may fail to accurately assign a variant to its biologically relevant gene, particularly when the variant is located in a region where multiple genes overlap. Our framework identified SNP loci that warrant further investigation in future research. These genetic markers could provide valuable information on the biological mechanisms underlying the susceptibility of disease in diverse ancestries and potentially serve as targets for therapeutic intervention.

## Methods

### Data Preprocessing

To reduce the dimensionality of the input data and mitigate the effect of LD, we processed the QC’ed SNPs using LD clumping, performed exclusively on the European training set to prevent data leakage. This procedure consisted of two steps. First, we conducted a standard GWAS using PLINK v2.00 to obtain the association between each cardiometabolic disease and each SNP. Second, we performed LD clumping using relatively inclusive thresholds (– clump-p1 0.05, –clump-p2 0.05, –clump-r2 0.7, –clump-kb 500) based on the *p*-values derived from the GWAS. This inclusive threshold was chosen to retain a larger number of potentially informative variants for the deep learning model. The LD clumping procedure selected a tag SNP with the smallest *p*-value within each LD group. As a result, each CMD had a unique list of variants. On average, the dimensionality of SNPs was reduced from 13,277,480 to approximately 100,000 for each disease, with the number of variants varying from 10,000 to 2,000 across chromosomes. This reduction in dimensionality not only simplifies the computational complexity of the analysis but also enables the model to focus on the most informative SNPs while accounting for the redundancy introduced by LD.

Based on the UK Biobank documentation, the CMDs were defined through multiple data sources. Atherosclerosis uses hospitalization records (ICD-9 414.0, 440.*; ICD-10 I70.*, I25.0, I25.1), GP Read codes, and death registry information. Atrial fibrillation incorporates self-reported codes (field 20 002), ICD-10 I4 8, and extensive GP Read codes. Cardiovascular disease was defined as a composite endpoint comprising three main conditions: coronary artery disease, peripheral vascular disease (ICD-9 443.8-44.9, ICD-10 I73.8-I73.9 and self-reported codes) and ischemic stroke (ICD-9 433.*, 434.*, ICD-10 I63.*, and self-reported cases). Coronary Artery Disease was defined using ICD-9 410.*-412.*, 414.0, 414.8, 414.9, ICD-10 I21.*-I24.*, I25.1, I25.2, I25.5, I25.6, I25.8, I25.9, and relevant coronary intervention procedures identified through OPCS codes (K40.*-K46.*, K49, K50.1, K50.2, K50.4, K 75.*). The definition was further enhanced by incorporating self-reported codes, surgical operation codes (field 20004 for PTCA/CABG/triplet-bypass procedures), and doctor-diagnosed health conditions (field 6150). Myocardial infarction uses doctor-diagnosed health conditions (heart attack), self-reported codes (heart attack), surgical operation codes, ICD-9 410.*-412.*, ICD-10 I21.*, I22.*, I24.1, I25.2, and the date of myocardial infarction (field 42000). Stroke uses ICD-9 430.*, 431.*, 434.*, 436.*, ICD-10, I60.*, I61.*, I63.*, I64.*, and self-reported codes. Type 2 diabetes was comprehensively defined through multiple data sources: self-reported diabetes (fields 20002, 2443, 4041), diabetes medication usage (fields 6153, 6177, 20003), laboratory values (HbA1c ≤ 6.5%, field 30750), and hospitalization records (ICD-9 code 250.* a nd ICD-10 codes E10.*-E14.*). Type 1 diabetes cases were specifically excluded from this definition to ensure proper phenotyping.

The dosage value of the minor allele for each SNP varies within the range of [0, 2], where 0 represents the absence of the minor allele, 1 represents the presence of one copy of the minor allele (heterozygous), and 2 represents the presence of two copies of the minor allele (homozygous). The value embedding was performed using a one-dimensional convolutional layer. The positional information of the SNPs on each chromosome was obtained and converted to absolute position encodings using sine and cosine functions for geometric progression, as proposed in the vanilla Transformer architecture [33]. Finally, together with the covariates, the input features were normalized using standard scaling to ensure that the covariates have zero mean and unit variance, which improves the convergence and stability of the deep learning model during training.

### Model Architecture of DeepGP

DeepGP is a multi-input deep learning model designed to estimate the risk probability of CMDs using input data consisting of SNPs and covariates. The model architecture incorporates 22 Bi-SSM branches, each processing the SNP data from one of the 22 autosomal chromosomes. This formulation allows for an effective capture of the temporal dependencies and interactions among the genetic variants along the chromosome. In addition, DeepGP includes a FCN module that processes the covariates, which allows the model to consider the contributions of environmental factors to the overall disease risk. This multi-input architecture enables DeepGP to effectively harness the information from multi-modal data and provide more accurate and personalized risk predictions for CMDs.

The Bi-SSM branches are specialized in capturing the complex, non-linear relationships between the genetic variants and the disease risk. On each branch, the first step is to process SNP dosages via an embedding layer to obtain the desired dimension (*N*) of hidden states and add positional encoding. The core idea is a structured state space sequence model that maps an input sequence *x*(*t*), i.e., embedding representation of SNP *x* on position *t* for the first layer, to an output sequence *y*(*t*) with an intermediate hidden state representation *h*. The model is constructed with system parameters **A, B**, and **C** as follows:

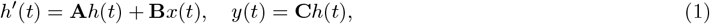

where **A** ∈ ℝ^*N* × *N*^ is the state matrix for evolution parameters, and **B** ∈ ℝ^*N* ×1^ and **C** ∈ ℝ^1× *N*^ represent the projection parameters. The state matrix **A** governs the transition dynamics of the hidden states, while the projection matrices **B** and **C** map the input sequence and hidden states to the appropriate dimensions. In practice, the discrete form of these continuous parameters can be denoted as follows with zero-order hold transformation

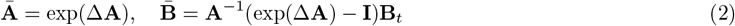

where Δ is a time scaler parameter, i.e., step size, and therefore Eq. (1) can rewritten as follows:

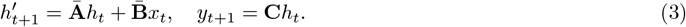

To enable the model to attend to specific inputs, Mamba introduces a selection mechanism that projects parameters **B** and **C** via linear transformations on *x*_*t*_, and uses Δ, which also linearly projects *x*_*t*_ via a linear transformation followed by a non-linear activation function. In the Bi-SSM module, we utilize two Mamba modules to process hidden states in both forward and backward directions and concatenate them together at the output. The SNP data are processed by two Bi-SSM modules. Combining the covariate representation *y*_*cov*_ from the FCN the output of DeepGP is denoted as follows:

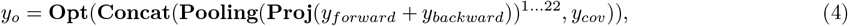

where **Opt** represents the output head consisting of a MHSA module followed by a FCN, **Concat** performs concatenation of features, **Proj** applies a projection transformation, and **Pooling** aggregates the projected bidirectional features from the combined forward (*y*_*forward*_) and backward (*y*_*backward*_) representations, which are then combined with covariate features (*y*_*cov*_). *y*_*o*_ is a risk probability processed by a sigmoid layer for binary classification.

### Model Training and Evaluation

To address the highly imbalanced ratio of cases to controls for the eight selected CMDs in the UK Biobank, which ranges from 2.5% (stroke) to 9.8% (coronary artery disease), we constructed a balanced development dataset for each disease by ensuring an equal number of cases and controls. This approach ensures that the model is trained on an equal representation of both classes, mitigating potential bias towards the majority class.

From the balanced development set, we randomly allocated 15% of the cases and controls to create a hold-out testing set for unbiased performance evaluation. The remaining samples were assigned to the training set. To prevent overfitting and optimize model hyperparameters, we employed a five-fold cross-validation scheme. The training set was divided into five equal-sized subsets, and in each fold, one subset was used as the validation set while the remaining four subsets were used for training. The hyperparameters that achieved the best average performance across the five folds, as measured by AUROC, were selected for the final model. Due to LD clumping, the selected SNP features in the input vary among the diseases. Therefore, the model validation was performed separately for each disease, with the hyperparameter ranges provided in Supplementary Table 4.

For classification tasks, we employed the binary cross-entropy loss function to optimize the model parameters, which measures the dissimilarity between the predicted probabilities and the true binary labels. To optimize the model parameters, we used the Adam optimizer, which adapts the learning rate for each parameter based on its historical gradients. Additionally, we incorporated early stopping and L2 regularization with a weight decay parameter in the Adam optimizer to further reduce the risk of overfitting.

All models in this study were implemented using Python 3.10. The deep learning models, including DeepGP, CNN and Dense shown in Supplementary Fig. 5, were developed using PyTorch 2.1.1. We leveraged PyTorch Lightning 2.0.8 to accelerate training and enable efficient utilization of multiple GPUs. All deep learning models were trained on NVIDIA A10 Tensor Core GPUs with CUDA 11.8. For machine learning approaches, we built LGBM models using LightGBM 4.3, while scikit-learn version 1.3.2 was used to implement PRS models using LR with L2 penalty, as well as RF, SNP-level LR, and MLP. To ensure fair comparison, all models received the same input data, including SNPs and covariates. Importantly, we applied identical cross-validation procedures to optimize multiple hyperparameters for all models using Optuna 3.6, maintaining methodological consistency across all comparisons. For instance, we optimized key hyperparameters in LGBM, including number of estimators, learning rate, maximum tree depth, and number of leaves.

### Phenotype Simulation

To evaluate the performance and robustness of DeepGP and the baseline methods, we conducted validation experiments using simulated phenotypes. These phenotypes were generated by a simulator that incorporates complex biological mechanisms, including intricate genetic architectures, gene-by-gene interactions (i.e., epistasis), and gene-by-environment interactions. This approach allows for a controlled assessment of the models’ ability to capture and predict complex trait outcomes under various scenarios that closely mimic real-world genetic complexity. Following the type 2 diabetes analysis setup, we used only European training and testing cohorts, incorporating SNPs from all autosomal chromosomes. We used the original genotype data from our preprocessing pipeline and sampled causal SNPs from a mixture of normal distributions, comprising 1% of the total number of SNPs, which reflects the complex genetic architecture of many traits. Given *N*_*s*_ sampled SNPs, the effect size variances were allocated as follows: large-effect SNPs 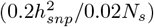, moderate-effect SNPs 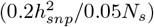, and small-effect SNPs 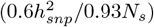 ensuring that 2%, 5%, and 93% of SNPs contribute to the total SNP heritability 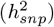, respectively [51].

We implemented a two-step process to incorporate complex genetic and environmental interactions. First, we randomly selected SNPs from the group with large effect sizes to model gene-by-gene interactions. For each selected SNP, we formulated a gene-by-gene interaction matrix and multiplied it with interaction effects, following an established approach for simulating epistatic effects [52]. Second, we introduced gene-by-environment interactions by modeling the target trait as a non-linear function of a heritable environmental factor [53]. The environmental factor was calculated as the product of the genotype matrix and a set of random SNP effects, plus residuals. This approach simulates a realistic scenario where genetic factors influence susceptibility to environmental exposures.

Finally, we rescaled the overall residuals to standardize the trait distribution. To generate binary traits, we transformed the continuous phenotypes into probabilities using a logistic sigmoid function. These probabilities were then used to sample from a binomial distribution, introducing an additional randomness to mimic the stochastic nature of disease occurrence. By incorporating these additional steps, we created a more realistic and challenging dataset for evaluating the predictive performance of DeepGP and baseline methods across a range of phenotypic distributions and outcome types.

### Interpretability Analysis

We employed DeepLIFT [44] to interpret the DeepGP model and identify the most important SNPs contributing to the risk predictions. DeepLIFT is a powerful feature attribution method that assigns importance scores to each input feature based on its contribution to the model’s output. This approach has gained significant attention and has been extensively applied in various domains of deep learning research, including gene discovery [54]. Moreover, it proves to be the most suitable option for our study due to its computational efficiency and memory-friendly nature, when compared with other interpretability analysis methods, such as SHAP (SHapley Additive exPlanations) [55], which required substantial memory allocation and led to out-of-memory issues on GPUs.

For a given input *x* and a reference input 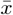, DeepLIFT calculates the importance score 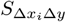 for each feature *i* as follows:

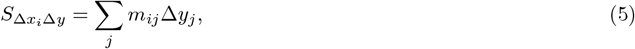

where *m*_*ij*_ is the multiplier that represents the contribution of the change in neuron *j* to the change in the output, and 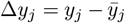 is the difference between the activation of neuron *j* and its reference value. The difference in input feature values is given by 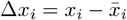.

The multipliers are computed recursively using a chain rule-like approach, allowing importance scores to be propagated back through the layers of the neural network:

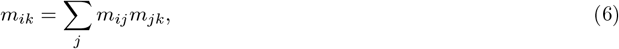

where *m*_*ik*_ represents the total contribution of the change in input feature *i* to the final output via all intermediate neurons. We applied DeepLIFT to the trained DeepGP model to obtain the importance scores for each SNP in the input data. The reference input was set to the mean value of each SNP across all samples. Since the SNP data were standardized, this reference value is zero, providing a consistent baseline for calculating importance scores. The resulting importance scores were then compared to the Chi-square statistics derived from our GWAS to assess the concordance between the model’s feature attributions and the statistical significance of the SNPs.

## Data availability

The UK Biobank is an open access resource available to approved researchers. Access to the data used in this study is available through an application procedure described at https://www.ukbiobank.ac.uk/enable-your-research.

## Code availability

The code implementation of DeepGP is available on GitHub (https://github.com/tndrg/DeepGP).

## Acknowledgements

This research has been conducted using the UK Biobank Resource under Application Number 53639. Taiyu Zhu is fully funded by Novo Nordisk and is a Novo Nordisk Postdoctoral Fellowship run in partnership with the University of Oxford. We would like to thank Neil Robertson at Novo Nordisk Research Centre Oxford for his contribution in data processing. We also thank the UK Biobank participants and the UK Biobank team for their work in collecting, processing, and disseminating these data for analysis.

## Supplementary

### Data Split

Table 3 presents the number of samples in the training and testing sets for each cardiometabolic disease, along with the total number of cases. These samples were obtained after filtering to include only unrelated European participants based on the UK Biobank’s quality control procedures. Disease classifications were derived using the multiple-source data definition approach within the UK Biobank’s defined disease phenotypes, which integrates information from ICD codes, self-reports, hospital records, and other clinical data sources to ensure robust phenotyping.

**Table 3:**
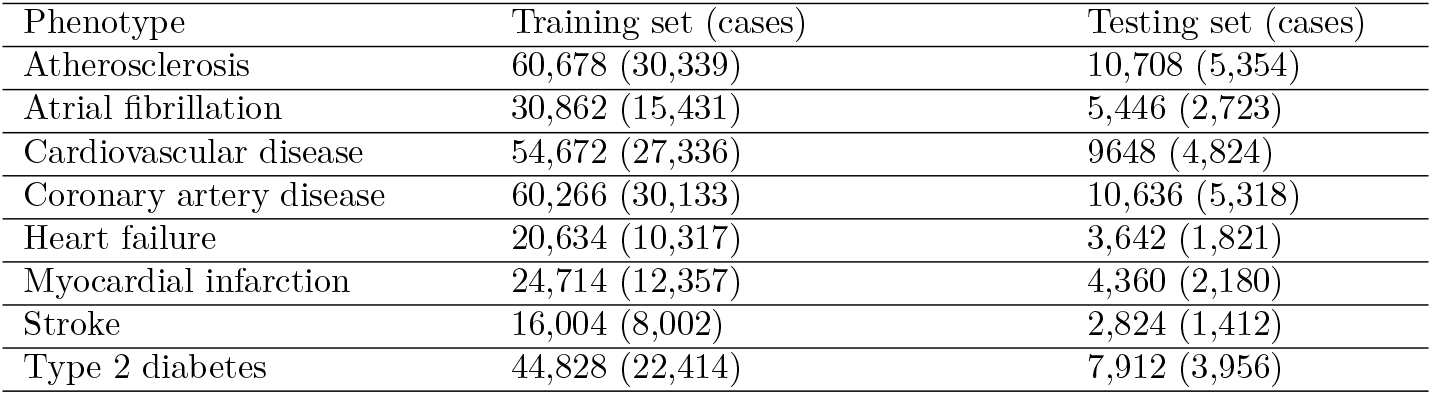
Sample sizes of the training and testing sets for the considered CMDs in the UK Biobank.

| Phenotype | Training set (cases) | Testing set (cases) |
| --- | --- | --- |
| Atherosclerosis | 60,678 (30,339) | 10,708 (5,354) |
| Atrial fibrillation | 30,862 (15,431) | 5,446 (2,723) |
| Cardiovascular disease | 54,672 (27,336) | 9648 (4,824) |
| Coronary artery disease | 60,266 (30,133) | 10,636 (5,318) |
| Heart failure | 20,634 (10,317) | 3,642 (1,821) |
| Myocardial infarction | 24,714 (12,357) | 4,360 (2,180) |
| Stroke | 16,004 (8,002) | 2,824 (1,412) |
| Type 2 diabetes | 44,828 (22,414) | 7,912 (3,956) |

### Model Details and Hyperparameters

Table 4 presents the hyperparameter ranges evaluated during model validation. For each of the considered CMDs, the final hyperparameter configuration was selected based on the best AUROC performance, resulting in slight variations in the optimal model parameters across different diseases. These disease-specific optimizations ensure that each model is tailored to capture the unique genetic architecture and risk factors associated with each particular condition.

**Table 4:**
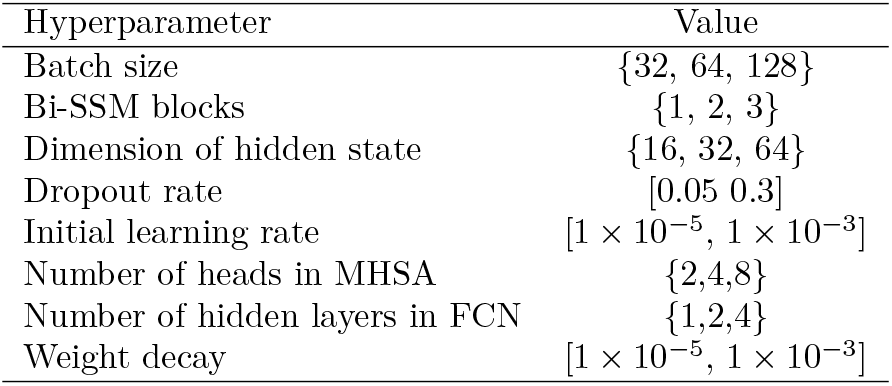
Ranges of hyperparameter tuning in model validation.

### Comparative Performance Analysis

Table 5 provides additional evaluation metrics for genomic risk prediction across eight CMDs in the European ancestry population, while Table 6 presents supplementary metrics for validation on African and Caribbean ancestries specifically for type 2 diabetes. Fig. 5 in Supplementary illustrates the AUROC curves of various traditional machine learning and deep learning methods in genomic risk prediction for the considered CMDs.

**Table 5:** Performance of genomic risk prediction on hold-out testing data of European ancestry.

| Phenotype | Method | Accuracy | Precision | Sensitivity | Specificity | AUROC |
| --- | --- | --- | --- | --- | --- | --- |
| Atherosclerosis | DeepGP | **68.6** | **67.1** | **72.8** | **64.3** | **0.75** |
|  | LGBM | 64.7 | 64.3 | 66.4 | 63.2 | 0.70 |
|  | PRS | 65.7 | 64.2 | 70.7 | 60.7 | 0.70 |
| Atrial fibrillation | DeepGP | **67.6** | **68.9** | 64.1 | **71.1** | **0.74** |
|  | LGBM | 65.7 | 65.0 | 68.2 | 63.3 | 0.71 |
|  | PRS | 65.9 | 64.1 | **72.2** | 59.6 | 0.70 |
| Cardiovascular disease | DeepGP | **67.1** | **67.5** | 65.9 | **68.3** | **0.74** |
|  | LGBM | 63.8 | 63.4 | 65.6 | 62.1 | 0.69 |
|  | PRS | 64.4 | 62.5 | **72.1** | 56.8 | 0.68 |
| Coronary artery disease | DeepGP | **67.4** | **67.5** | 66.9 | **67.8** | **0.74** |
|  | LGBM | 63.9 | 63.4 | 65.8 | 62.0 | 0.68 |
|  | PRS | 63.5 | 62.0 | **69.9** | 57.2 | 0.69 |
| Heart failure | DeepGP | **67.1** | **64.0** | **78.3** | 55.9 | **0.72** |
|  | LGBM | 62.8 | 62.8 | 62.6 | **62.9** | 0.68 |
|  | PRS | 62.7 | 62.0 | 66.3 | 59.2 | 0.66 |
| Myocardial infarction | DeepGP | **69.0** | **72.0** | 62.3 | **75.7** | **0.76** |
|  | LGBM | 65.8 | 65.1 | 68.2 | 63.5 | 0.71 |
|  | PRS | 62.9 | 61.6 | **68.3** | 57.3 | 0.67 |
| Stroke | DeepGP | **63.2** | **62.0** | **68.2** | **58.3** | **0.68** |
|  | LGBM | 59.6 | 59.5 | 59.8 | 59.3 | 0.62 |
|  | PRS | 57.9 | 57.5 | 60.8 | 55.1 | 0.60 |
| Type 2 diabetes | DeepGP | **60.4** | **59.5** | **65.5** | 55.5 | **0.64** |
|  | LGBM | 59.5 | 59.2 | 59.6 | **58.9** | 0.61 |
|  | PRS | 57.7 | 57.4 | 59.8 | 55.7 | 0.60 |

**Table 6:** Performance of genomic risk prediction for type 2 diabetes on African and Caribbean ancestries.

| Phenotype | Method | Accuracy | Precision | Sensitivity | Specificity | AUROC |
| --- | --- | --- | --- | --- | --- | --- |
| DeepGP | African | **76.9** | **31.5** | 32.3 | **85.9** | **0.64** |
|  | Caribbean | **76.1** | **34.3** | 37.4 | **84.5** | **0.68** |
| LGBM | African | 53.7 | 18.9 | **53.6** | 53.7 | 0.54 |
|  | Caribbean | 54.9 | 19.8 | **50.3** | 55.9 | 0.56 |

**Fig. 5:**
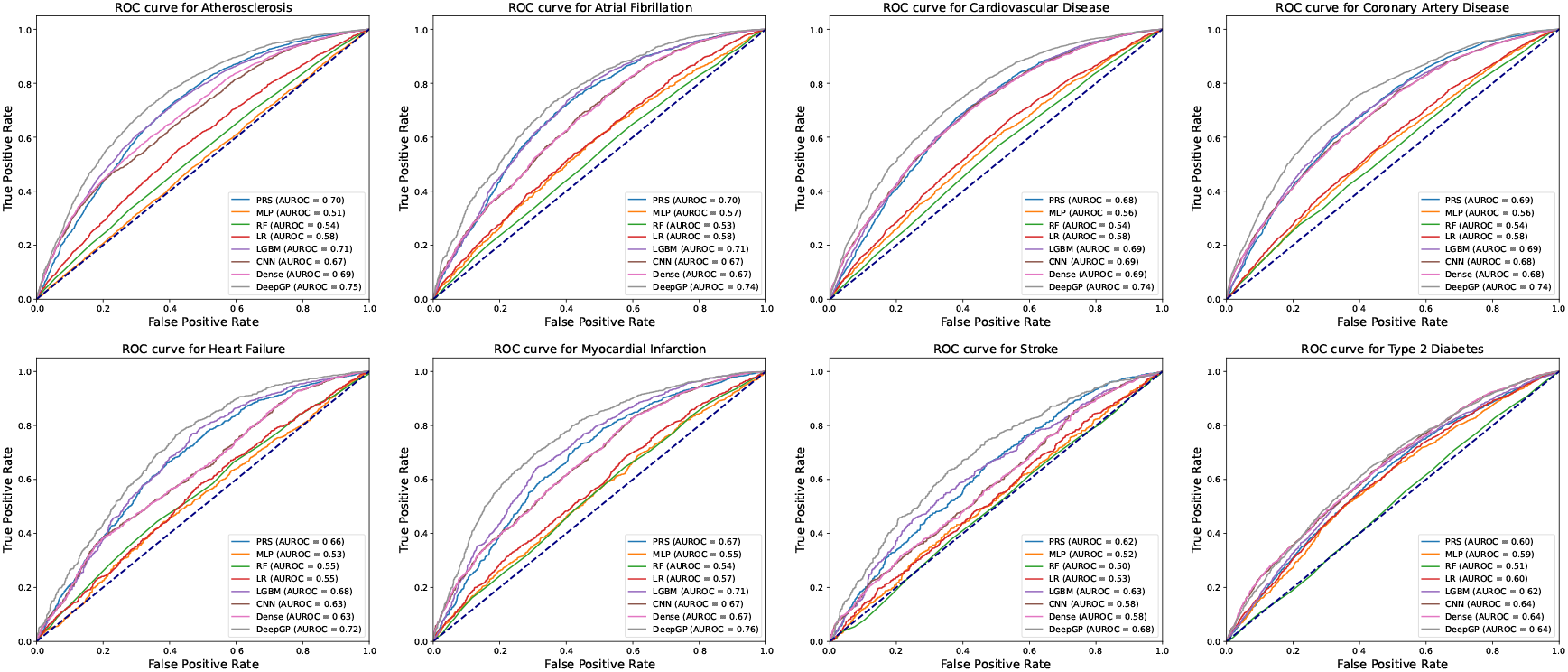
Predictive performance across all the considered methods. AUROC for DeepGP and baseline methods: PRS, multilayer perceptron (MLP), random forest (RF), logistic regression (LR), LGBM, convolutional neural network (CNN), and dense neural network (Dense).

